# Complementary Use of Resveratrol Improves Low-Grade Chronic Inflammation Unresolved by Standard DMARD Therapy in Rheumatoid Arthritis

**DOI:** 10.64898/2026.07.24.26358846

**Authors:** Aharna Guin, Sanchaita Misra, Dipanjan Bhattacharya, Sudipta Chatterjee, Alakendu Ghosh

## Abstract

**Background:** Chronic low-grade inflammation in long-standing rheumatoid arthritis (RA) contributes not only to joint damage but also to metabolic dysregulation, endothelial dysfunction, and elevated cardiovascular (CV) risk. Although combination disease-modifying anti-rheumatic drugs (DMARDs) remain the mainstay of therapy, their long-term efficacy in controlling systemic inflammation and preventing metabolic complications appears limited. Phytochemicals such as **resveratrol**, a polyphenolic compound widely used in traditional and complementary medicine possess anti-inflammatory and immunomodulatory properties.

**Objectives:** To investigate whether resveratrol can complement the immunomodulatory effects of combination DMARDs in long-duration RA patients by modulating inflammatory cytokines and the JNK–IRS–Akt insulin signaling axis.

**Methods:** This study enrolled early and late rheumatoid arthritis patients to assess disease activity, vascular markers, and ex-vivo PBMC responses. PBMCs were isolated for cytotoxicity testing and resveratrol treatment, followed by ELISA and Western blot analysis. Statistical comparisons evaluated immunomodulatory effects and inflammatory signaling alterations.

**Results:** Longitudinal follow-up of RA patients showed significant first-year improvement in disease activity and atherosclerotic markers, correlated with MTX dose. Early versus late RA comparison revealed heightened cytokines and enhanced JNK-mediated stress signaling in longstanding disease. Resveratrol displayed safe PBMC viability, reduced LPS-induced TNF-α and adipokines, and downregulated p-JNK and GSK-3β, indicating targeted anti-inflammatory modulation independent of Akt activation.

**Conclusion:** Chronic RA showed persistent inflammatory and metabolic dysregulation driven by JNK–NF-κB activation. Resveratrol reduced cytokines, corrected adipokine imbalance, and selectively inhibited JNK, suggesting adjunct therapeutic value alongside DMARDs for improving immunometabolic disturbances in long-standing RA.

**Graphical Abstract:** 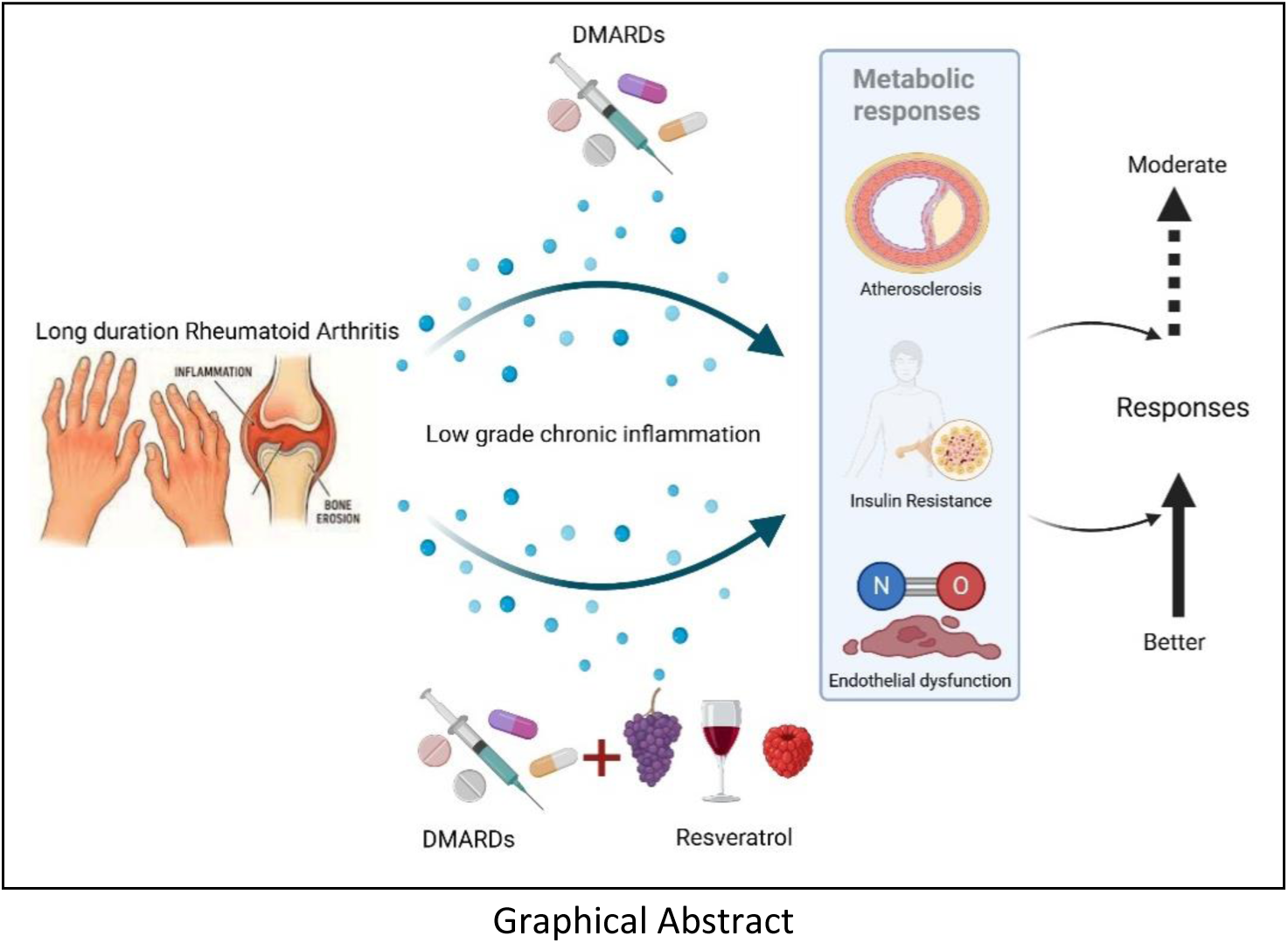

*Highlights of the study:*

- DMARDs therapy against atherosclerotic indices tends to plateau in long-term.
- Chronic RA elevates TNF-α, leptin levels and activates JNK–IRS-1–Akt signaling.
- Resveratrol significantly downregulates cytokines and p-JNK signaling in PBMCs.

## Introduction

Rheumatoid arthritis (RA) is a chronic systemic autoimmune disease characterized by persistent synovial inflammation, progressive joint destruction, and significant extra-articular manifestations [1, 2]. Over the past several decades, therapeutic strategies for RA have undergone substantial evolution—from early reliance on corticosteroids to the widespread adoption of combination conventional synthetic disease-modifying antirheumatic drugs (cDMARDs), and subsequently to biologics and targeted small-molecule therapies [3,4]. Despite these advancements, **cDMARDs remain the most extensively used globally owing to their accessibility, affordability, and established clinical efficacy [5]**.

Conventional DMARDs, particularly methotrexate (MTX), remain the cornerstone of RA management and are highly effective in reducing disease activity and retarding radiographic progression **[6]**. Clinical evidence strongly supports that **early diagnosis and early initiation of DMARD therapy**—preferably within the first year of disease onset—provides optimal control of inflammation and improves long-term outcomes **[7]**.

Beyond their role in joint preservation, DMARDs may also influence systemic complications of RA. Cardiovascular diseases (CVD) constitute the leading cause of mortality in RA patients **[8]**, largely driven by chronic inflammation, endothelial dysfunction, metabolic imbalance, and accelerated atherosclerosis. Several studies suggest that MTX and other DMARDs possess cardioprotective effects, either by modulating traditional atherosclerotic risk factors or by suppressing systemic inflammation that contributes to vascular damage **[9]**. However, the literature assessing DMARD-mediated cardiovascular benefits remains limited, and most studies have focused primarily on MTX rather than on combination therapies **[10]**.

**Chronic low-grade inflammation serves as a central mechanism in the etiopathogenesis and progression of RA** [11, 12]. Proinflammatory cytokines, chemokines, and altered adipokine secretion contribute not only to synovial proliferation and cartilage destruction but also to systemic metabolic disturbances **[13]**. Elevated TNF-α levels activate intracellular stress pathways that disrupt insulin receptor substrate-1 (IRS-1) function, impair the PI3K-dependent signaling cascade, and thereby promote insulin resistance (IR) **[14]**. Notably, early aggressive treatment has been shown to reduce both inflammatory activity and the rate of atherosclerotic progression within the first year of diagnosis **[15, 16]**.

Despite the therapeutic value of DMARDs, **adverse effects, incomplete responses in chronic disease, and the high cost of biologics, particularly in low-resource settings, frequently limit long-term use [17, 18]**. These limitations have generated growing interest in complementary and plant-derived therapeutic agents [19]. Phytochemicals—such as **resveratrol, curcumin, and epigallocatechin gallate**—are widely recognized for their anti-inflammatory, antioxidant, and immunomodulatory properties, [20, 21] and have demonstrated beneficial effects in various chronic conditions including cancers, neurodegenerative disorders, and autoimmune diseases [22, 23, 24]. Importantly, several of these compounds have been shown to modulate key signaling pathways involved in RA pathogenesis, including the NF-κB, MAPK, and PI3K-Akt axes [25, 26, 27].

Given the **diminishing long-term effectiveness of combination DMARDs in controlling chronic low-grade inflammation** and the growing evidence supporting the therapeutic potential of phytochemicals, this study aimed to investigate the **ex-vivo effects of resveratrol**, a well-characterized plant-derived polyphenol, on peripheral blood mononuclear cells (PBMCs) from long-standing RA patients.

## Materials and Methods

### Study Participants and Sample Collection

Patients with rheumatoid arthritis (RA), aged 18–60 years, who fulfilled the American College of Rheumatology (ACR) 1987 classification criteria for RA [28], were recruited from the Outpatient Department of Rheumatology, Institute of Postgraduate Medical Education and Research (IPGME&R), SSKM Hospital, Kolkata. Individuals with a prior history of coronary artery disease, cerebrovascular accident, smoking, or co-morbid disorders including diabetes mellitus, obesity (BMI ≥30 kg/m²), familial dyslipidemia, peripheral vascular disease, hypothyroidism, renal impairment (serum creatinine ≥3.0 mg/dL or creatinine clearance ≤30 mL/min), liver disease, or Cushing’s syndrome were excluded. Patients receiving medications known to influence lipid metabolism (lipid-lowering agents, β-blockers, oral contraceptives, estrogens, progestins, thyroxine, and vitamin E) were also excluded. Pregnant or recently pregnant (within 3 months) women were not eligible.

Written informed consent was taken from all study participants. The study protocol had been approved by the institutional ethics committee of IPGME&R, Kolkata and have been performed in accordance with the ethical standards as laid down in ICMR Ethical Guidelines 2006.

### Study Design

Thirty patients with early RA (disease duration < 1 year) were enrolled for prospective follow-up and evaluated at baseline, 1 year, and 2 years. Disease activity was assessed using the Disease Activity Score-28 (DAS28) [29]. All patients received Methotrexate (7.5–20 mg/week) in combination with Hydroxychloroquine (200–400 mg/day) and Sulfasalazine (0.5–2 g/day), titrated according to disease activity. Short-term low-dose corticosteroids (≤7.5 mg/day for <3 months) were used when clinically necessary. Patients were reviewed every 3 months to document symptoms, clinical findings, dose adjustments, and lifestyle or dietary modifications. Body weight and BMI were recorded at each visit. High-sensitivity C-reactive protein (hsCRP) was measured by nephelometry (BN ProSpec, Siemens). Flow-mediated vasodilatation and carotid intima-media thickness were assessed following previously standardized methodology [16].

### Statistical Analysis

Data distribution was tested using the Shapiro–Wilk test. Variables are presented as mean ± standard error (SE) for comparability. Between-group differences were analyzed using one-way ANOVA with Bonferroni post-hoc correction for normally distributed data or Dunn’s multiple comparison test for non-parametric data. Independent-sample t-tests or Mann–Whitney U tests were used for two-group comparisons. A *p* value <0.05 was considered statistically significant. Analyses were performed using GraphPad Prism (version 9.0).

### PBMCs (peripheral blood mononuclear cells) Isolation

For ex-vivo studies, 20 early-stage (disease duration < 1 year) and 20 late-stage RA (disease duration > 5 years) patients were included. After overnight fasting (12 hours), venous blood was collected and peripheral blood mononuclear cells (PBMCs)/monocytes were isolated using HiSep LSM 1073 (Himedia, Mumbai) according to manufacturer instructions. Briefly, whole blood was layered over an equal volume of LSM-1073 and centrifuged at 3000 rpm for 40 minutes at room temperature. The PBMC/monocyte layer was aspirated, washed twice with phosphate-buffered saline (PBS, pH 7.2), and processed for downstream assays. Plasma fractions were collected and stored at −80 °C for biomarker estimation to be done within one month.

### Cytotoxicity Assay for Resveratrol Dose Determination

Cell viability following resveratrol treatment was evaluated using the 3-(4,5-Dimethylthiazol-2-yl)- 2,5-diphenyltetrazolium bromide (MTT) assay, as described by Mosmann (1983). PBMCs (5 × 10⁴ cells/well) were cultured in 96-well plates with increasing concentrations of resveratrol (R5010, Sigma) (500 ng/mL– 24 µg/mL) for 24 h. Untreated cells served as controls. After incubation, 20 µL of MTT solution (5 mg/mL) was added to each well and incubated for 4 h at 37 °C. Formazan crystals were dissolved in 100 µL DMSO, and absorbance was measured at 570 nm.

### Cell viability (%) = (ODₜᵣₑₐₜₑd / ODcₒₙₜᵣₒₗ) × 100 Resveratrol Treatment of PBMCs

PBMCs were washed in incomplete RPMI-1640 medium, counted using a hemocytometer, and seeded at 2 × 10⁶ cells/mL in complete medium supplemented with autologous serum. After a 1-h adherence phase at 37 °C in a CO₂ incubator, cells were treated with resveratrol doses of 10 µg/mL and 15 µg/mL each, followed by LPS stimulation (1 µg/mL) for 18 h. Supernatants were collected, centrifuged (3000 rpm, 4 minutes), and stored for ELISA. Cell pellets were used for Western blotting.

### Measurement of Pro-Inflammatory Cytokines and Adipokines

TNF-α and adipokines (adiponectin, leptin, resistin) were quantified from culture supernatants using RayBio® Human ELISA Kits (Cat. Nos. ELH-TNFa-1, ELH-Adiponectin-1, ELH-Leptin-1, ELH-Resistin-1) respectively, following the manufacturer’s protocol provided.

### Western Blot Analysis

PBMCs (2 × 10⁶ cells/mL) were lysed using 2× cell lysis buffer (Cell Signaling Technology, Beverly, MA) supplemented with protease and phosphatase inhibitors. Total protein was quantified using the Bradford assay. Equal quantities of protein (30–40 µg) were denatured with 5X loading buffer, resolved on 10% SDS-polyacrylamide gels, and electro transferred to nitrocellulose membranes. Membranes were blocked with 5% BSA for 1 h and incubated overnight with primary antibodies (1:1000), followed by HRP-conjugated secondary antibodies (1:1000). Immunoreactive bands were visualized using Novex® ECL Chemiluminescent Substrate (Invitrogen, USA) and documented with a Chemidoc DNR system. α-Tubulin served as the loading control. .

All cell culture reagents were procured from Gibco and Thermo life sciences Pvt Ltd. All antibodies were procured from Cell Signaling Technology. The resveratrol was procured from Sigma-Aldrich (R5010).

## Results

### Follow-up data of the RA cohort

A longitudinal evaluation of 30 RA patients over 1 year and subsequently 2 years demonstrated a significant reduction in disease activity and atherosclerotic indices after the first year of therapy, with minimal additional improvement during the second year (Figure 1).

**Figure 1:**
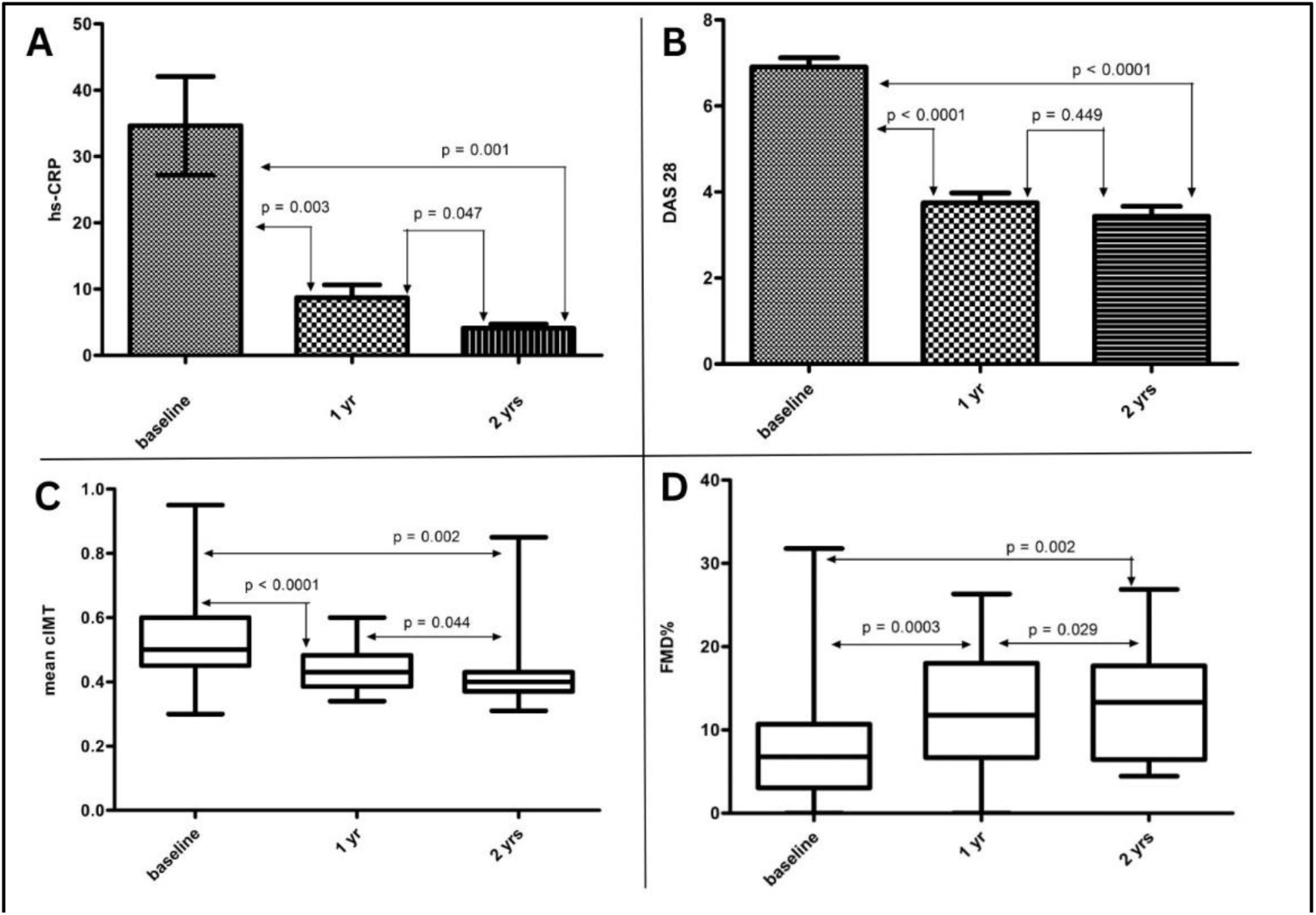
Follow-up effect of DMARDs on the disease activity and atherosclerotic indices of RA patients at base line, after 1 year and 2 years respectively

Analysis of drug exposure revealed that the mean weekly dose of methotrexate (MTX) showed a significant correlation with changes in both DAS28 scores and carotid intima–media thickness (cIMT) at the end of year 1. This correlation persisted with a similar trend at year 2 (Table 1).

**Table 1:** Correlation between avg. drug doses and changes in variables in the 1^st^ year & 2^nd^ year.

| Drugs | CHANGES (correlation, r) |  |  |  |  |  |  |  |  |  |
| --- | --- | --- | --- | --- | --- | --- | --- | --- | --- | --- |
|  | IMT |  | FMD |  | DAS 28 |  | hsCRP |  | ESR |  |
|  | 1<sup>st</sup> yr | 2<sup>nd</sup> yr | 1<sup>st</sup> yr | 2<sup>nd</sup> yr | 1<sup>st</sup> yr | 2<sup>nd</sup> yr | 1<sup>st</sup> yr | 2<sup>nd</sup> yr | 1<sup>st</sup> yr | 2<sup>nd</sup> yr |
| MTX | <u>0.425*</u> | <u>0.360<sup>†</sup></u> | 0.151 | -0.219 | <u>0.519**</u> | <u>0.349<sup>††</sup></u> | 0.095 | 0.083 | 0.213 | 0.081 |
| SSZ | -0.115 | 0.008 | -0.241 | -0.278 | -0.209 | 0.161 | 0.132 | -0.242 | 0.009 | 0.221 |
| HCQ | -0.038 | 0.049 | -0.123 | -0.341 | -0.229 | -0.018 | -0.308 | -0.175 | -0.211 | -0.037 |

\* p value = 0.015; \*\*p value = 0.003; <sup>†</sup> p value = 0.065; <sup>††</sup> p value = 0.075

### Comparison between Early and Late RA Patients

Pro-inflammatory mediators exhibited distinct patterns between early-stage and long-duration RA. Levels of TNF-α, resistin, and leptin were highest in long-duration RA, followed by early RA, and lowest in healthy controls (Figure 2). In contrast, adiponectin, an anti-inflammatory adipokine, showed a significant differential expression between early and late RA groups.

**Figure 2:**
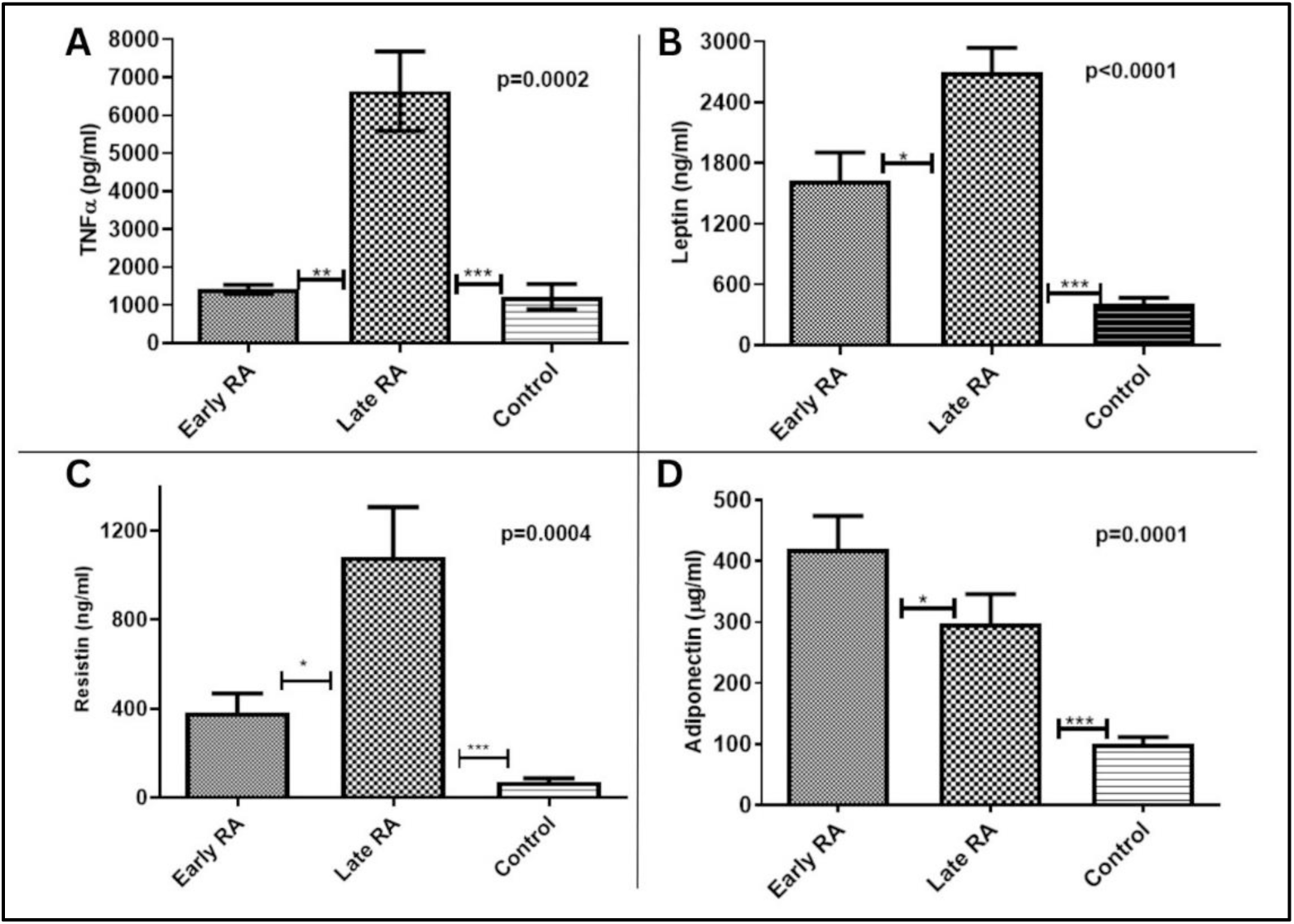
Comparative expression levels of different pro-inflammatory cytokines and adipokines among Early RA, Late RA and controls

Western blot analysis (Figure 3) demonstrated that phosphorylated JNK (p-JNK), phosphorylated IκBα (p-IκBα), and IRS-1 serine phosphorylation (IRS1-Ser^p) were markedly upregulated in long-duration RA. Conversely, tyrosine-phosphorylated IRS-1 (IRS1-Tyr^p), phosphorylated Akt (p-Akt), and GSK-3β were more prominently expressed in early RA. These findings indicate that chronic inflammatory burden enhances JNK-mediated stress signaling, alters PI3K–IRS-1 phosphorylation balance, and reduces Akt activation, collectively contributing to insulin-resistance-associated signaling impairment.

**Figure 3:**
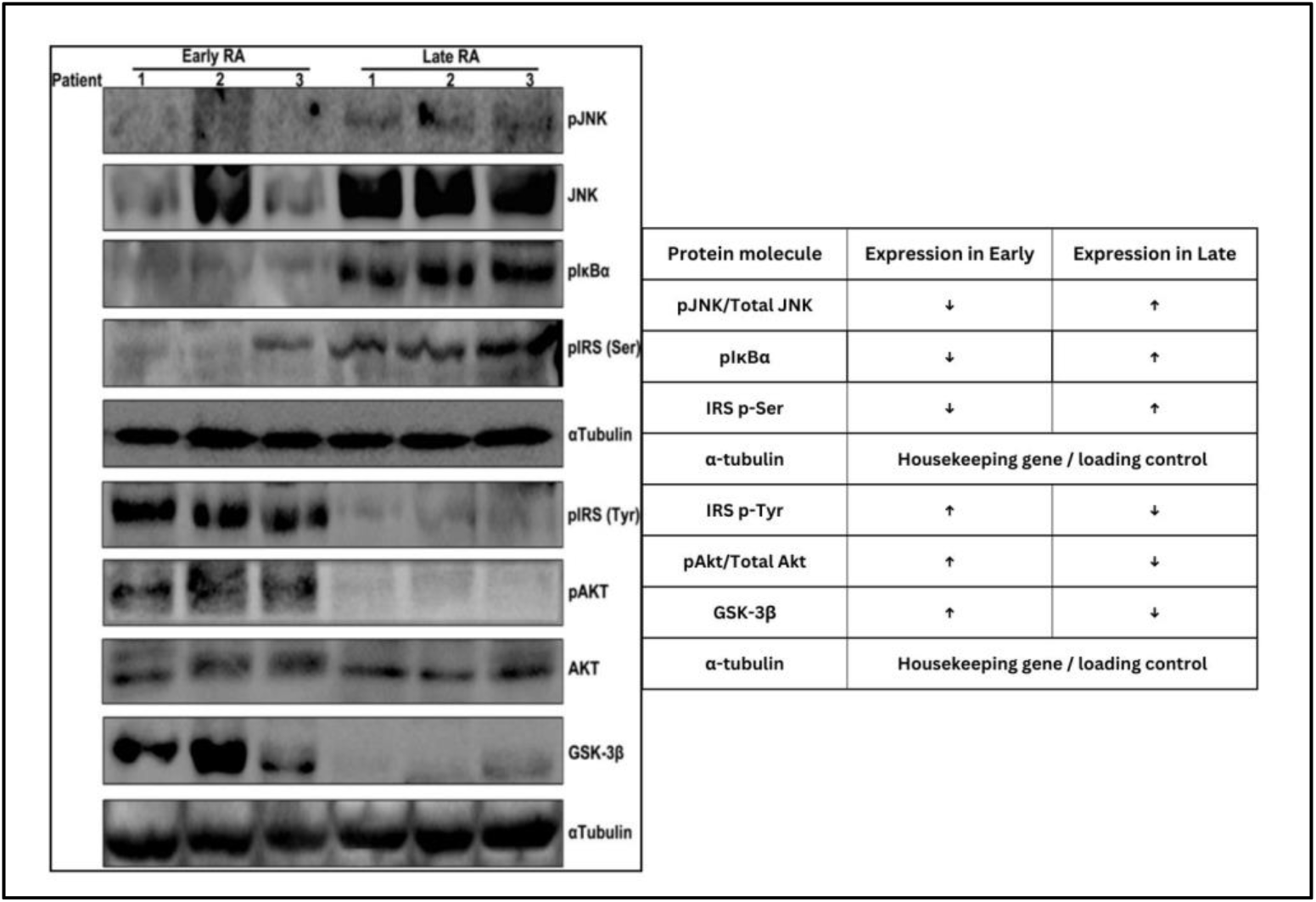
The expression levels of the different signalling molecules in the JNK-IRS-Akt mediated pathway of insulin action that probably is activated in RA inflamed tissues. JNK c-Jun N-terminal kinase; pIκBα-inhibitor of nuclear factor kappa-B kinase subunit alpha; IRS-1 Insulin receptor ubstrate-1; Ser Serine; Tyr Tyrosine; P phosphorylation; p-Akt phospho Protein kinase B; GSK-3β – glycogen synthetase kinase-3 subunit β.(↑) denotes upregulation; (↓) denotes downregulation.

### Cytotoxicity and Dose Selection of Resveratrol

Resveratrol, a phytochemical with documented anti-inflammatory potential, was evaluated as an adjunct therapeutic candidate. MTT assay revealed the IC₅₀ value was at 12.95μg/mL resveratrol (Figure 4). Based on this IC_50_ value, the lower and upper concentrations of **10μg/mL and 15μg/mL** were selected for mechanistic experiments.

**Figure 4:**
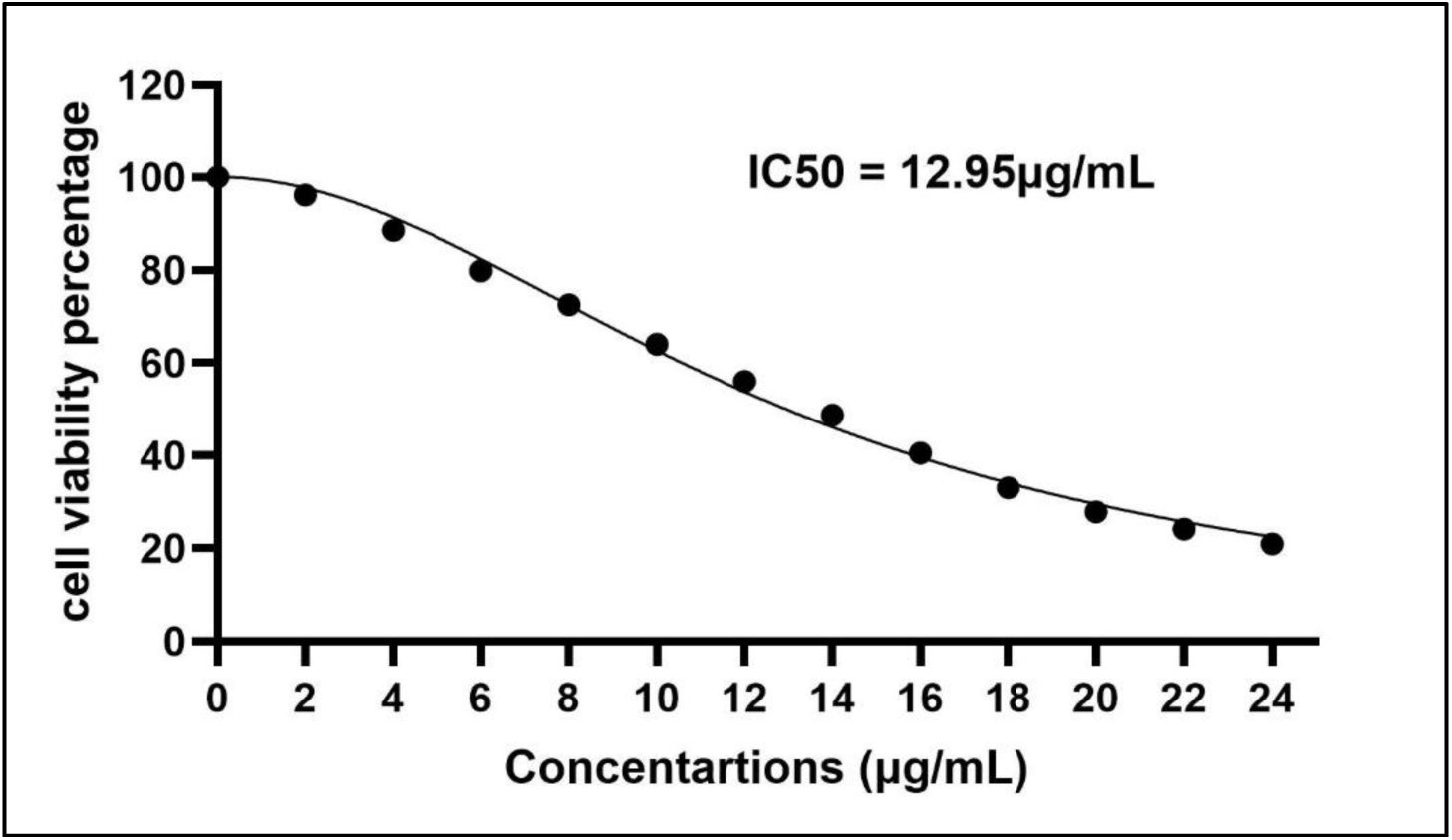
Cell viability assay of different doses of resveratrol.

### Effect of Resveratrol on Pro-inflammatory Molecules

Resveratrol significantly attenuated LPS-induced cytokine release (Figure 5a). Treatment with both 10 µM and 15 µM concentrations resulted in a marked reduction in TNF-α and adipokines secretion compared with the LPS-stimulated untreated group. The maximal inhibitory effect was observed at **10 µM**, indicating superior efficacy at this dose.

**Figure 5a:**
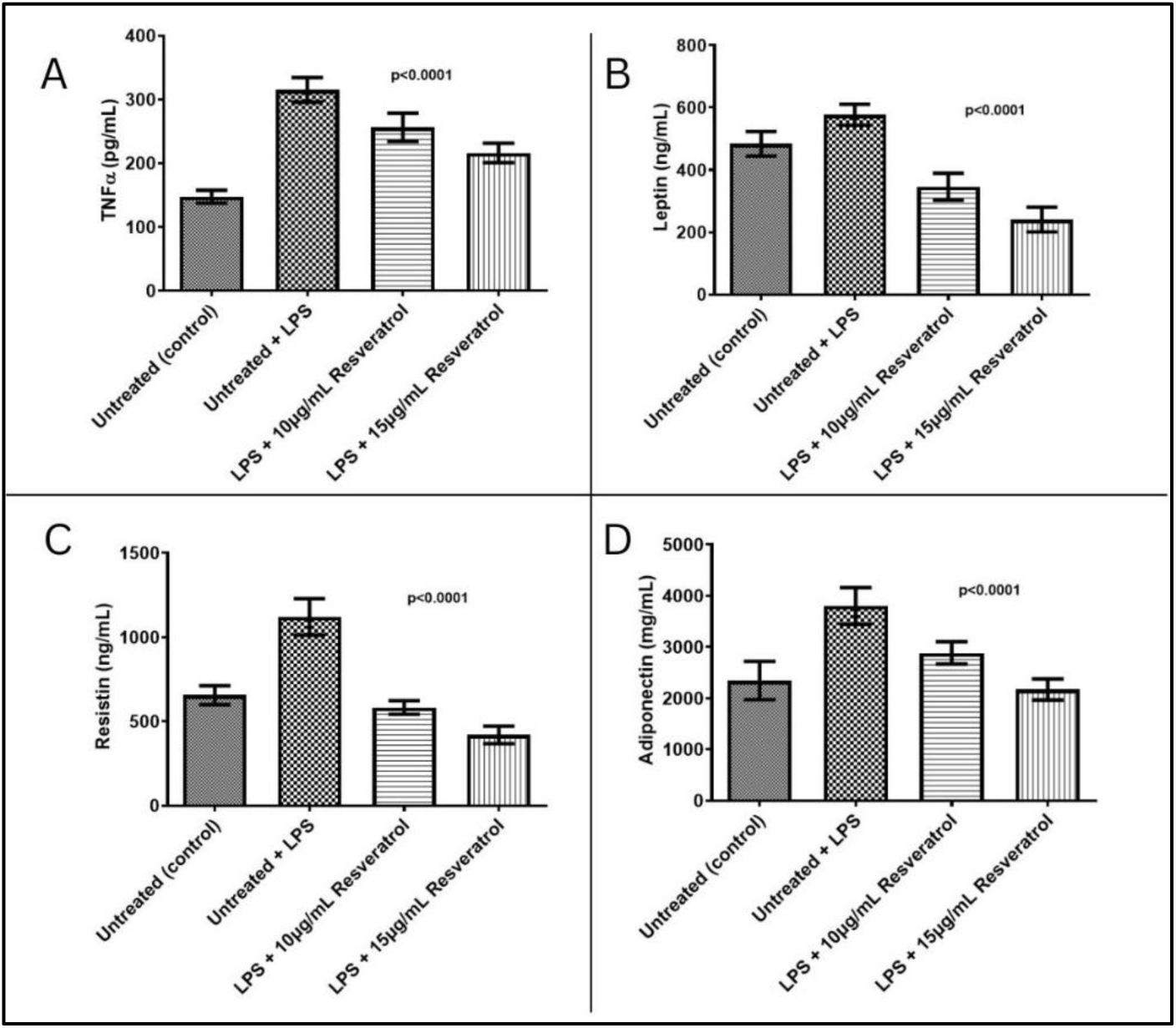
Effect of resveratrol on pro-inflammatory cytokines and adipokines.

**Figure 5b:**
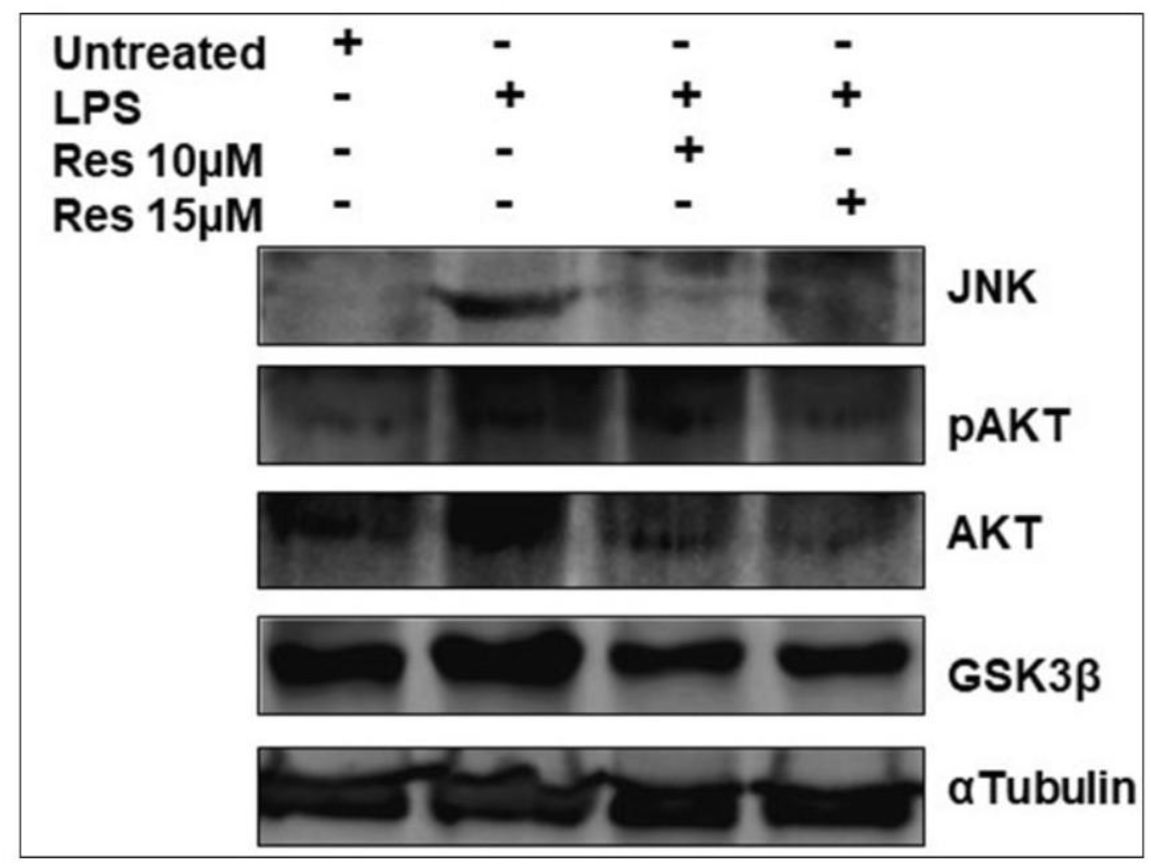
Effect of resveratrol on expression of JNK-Akt-GSK3β signaling molecules.

### Effect of Resveratrol on JNK–Akt Pathway Components

Resveratrol modulated key signaling intermediates of the JNK–Akt axis (Figure 5b). Treatment with 10 µM resveratrol produced the most pronounced reduction in p-JNK expression. GSK-3β levels were moderately reduced following treatment compared with LPS-stimulated cells. However, no appreciable change was observed in Akt or p-Akt expression following resveratrol exposure, suggesting that its principal mechanistic action in this context is mediated through downregulation of JNK and partial modulation of downstream effectors.

## Discussion

Rheumatoid arthritis (RA) is characterized not only by progressive joint destruction but also by the presence of multiple extra-articular comorbidities that significantly compromise long-term quality of life and survival [30]. Among these, metabolic disturbances and accelerated atherosclerosis have increasingly gained attention as integral components of chronic RA pathophysiology [31] In the present study**, alterations in key intracellular signaling molecules demonstrated a clear mechanistic link between RA-associated inflammation, cellular stress responses, and the development of insulin resistance** (IR). The proposed pathway (Figure 6) supports existing evidence that chronic inflammatory mediators serve as upstream triggers for metabolic dysregulation, even in the absence of obesity [32, 33].

**Figure 6:**
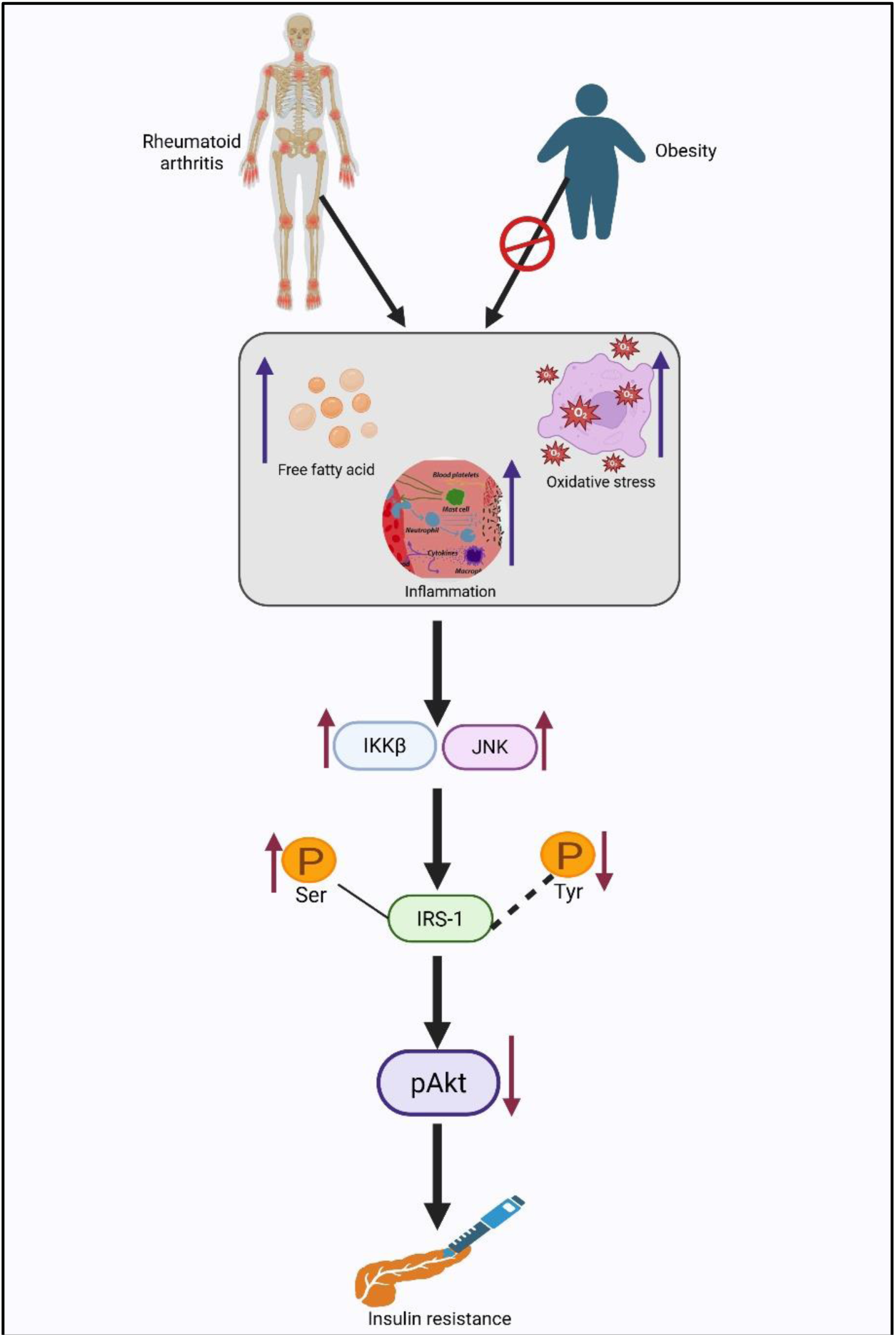
The probable signalling pathway connecting rheumatoid arthritis with insulin resistance.

Under normal physiological conditions, obesity-related free fatty acids (FFAs) and oxidative stress induce activation of stress kinases such as c-Jun N-terminal kinase (JNK) and inhibitor of κB kinase (IKK), which subsequently favor serine phosphorylation of IRS-1 over the canonical tyrosine phosphorylation, ultimately reducing downstream Akt/PKB activation and contributing to IR [34,35,36]. Our findings suggest that a similar cascade is initiated in chronic RA, wherein persistent elevations of TNF-α and other pro-inflammatory cytokines and adipokines mimic obesity-induced metabolic stress. **This provides a plausible biochemical rationale for the metabolic and vascular complications frequently observed in long-standing RA**.

The follow-up cohort analysis further revealed that methotrexate (MTX), a cornerstone DMARD, exerts anti-atherogenic effects that may extend beyond its anti-inflammatory actions. The observed reduction in cIMT after one year of MTX therapy is consistent with preclinical reports demonstrating reduced apoptotic cell accumulation and attenuated release of inflammatory mediators following MTX administration in murine models [37, 38]. However, **our data indicate that after two years, MTX, either alone or in combination with other conventional DMARDs, did not maintain its initial protective effect on inflammatory or atherosclerotic indices**. This plateau in therapeutic benefit underscores the need for adjunctive strategies that can sustainably modulate chronic inflammation and associated metabolic derangements.

Growing global interest in plant-derived therapeutic compounds has led to renewed focus on phytochemicals with immunomodulatory, antioxidant, and anti-diabetic properties [39]. Among these, resveratrol, quercetin, and stevioside have shown significant promise across inflammatory and metabolic disorders. Resveratrol, in particular, is well recognized for its ability to attenuate oxidative stress and inhibit the expression of pro-inflammatory cytokines, cyclooxygenases, protein kinases, and matrix metalloproteinases [40]. It’s chemopreventive and cytoprotective effects have been linked to modulation of PI3K-dependent glucose metabolism and the suppressive regulation of Akt/PKB phosphorylation [41]. Experimental studies in hepatocytes report that resveratrol inhibits insulin-stimulated interactions between IRS-1 and PI3K, as well as between IRS-1 and Grb2, thereby altering MAPK and PKB/Akt signaling cascades [42]. Additionally, resveratrol has been shown to suppress inducible nitric oxide synthase (iNOS) expression, decrease nitric oxide (NO) production, and reduce secretion of TNF-α and resistin in Kupffer cells and adipocytes exposed to LPS or hyperglycemic stress [43].

Consistent with these mechanistic insights, our study demonstrates **that resveratrol significantly down-regulated pro-inflammatory cytokines and adipokines—particularly leptin and resistin—in PBMCs derived from long-duration RA patients**. While several experimental models have reported anti-adipokine effects of resveratrol in obesity and metabolic syndrome, to our knowledge, this is among the earliest studies to document similar effects within an autoimmune disease context. Only a single randomized controlled trial by Khojah et al. (2018) has evaluated resveratrol supplementation in RA patients; however, the study involved oral administration, unlike our ex vivo mechanistic evaluation [44]. Our observations suggest that **chronic RA engages obesity-like inflammatory pathways, which are responsive to targeted modulation by resveratrol**.

Furthermore, resveratrol at 10µM substantially attenuated the PI3K–IRS-1–JNK signaling perturbations observed in late RA patients, indicating its potential to alleviate inflammation-driven metabolic stress. While its role in inhibiting the PI3K–Akt/mTOR axis has been well established in oncology research, the present findings extend its relevance to autoimmune pathology. Although, a recent review [45] has highlighted various signalling pathways via which resveratrol can affect RA pathology but the PI3K–IRS-1–JNK signaling axis has not been reported as such. ***This study could be one of the few reporting the PI-3-K insulin pathway as the target of resveratrol in long standing chronic inflammatory ex-vivo model*.**

Given the long-term nature of autoimmune diseases and the substantial side-effect burden associated with prolonged DMARD or biologic use, [46] **incorporating safe, cost-effective phytochemical adjuncts may help mitigate disease chronicity and reduce financial strain on patients** [47]. By dampening chronic inflammatory signaling and restoring portions of the metabolic axis, **resveratrol may serve as a useful adjunct to existing DMARD regimens, particularly in long-standing RA where drug response diminishes over time.**

The findings of this study underscore the therapeutic promise of resveratrol as a complementary agent in RA management. However, larger clinical studies with extended follow-up are warranted to validate these observations and to explore the translational potential of resveratrol in RA and other autoimmune conditions.

## Conclusion

In conclusion, the present study demonstrates that chronic low-grade inflammation in long-standing rheumatoid arthritis is closely linked to dysregulation of the JNK–IRS-1–Akt signaling axis and insulin-resistance–associated pathways. Although conventional DMARD therapy, particularly methotrexate, significantly reduces disease activity and atherosclerotic indices during early treatment, its long-term benefits appear to plateau. Ex vivo findings reveal that resveratrol effectively attenuates pro-inflammatory cytokines and adipokines and downregulates stress kinase signaling, particularly p-JNK, in PBMCs derived from RA patients. **These results highlight the potential of resveratrol as a complementary adjunct to standard DMARD therapy for mitigating persistent inflammation and metabolic dysfunction in chronic RA.** Further large-scale clinical studies are warranted to validate its translational applicability in routine RA management.

## Supporting information

the original blots for western blot data of fig 3

the original blots for western blot for figure 5b

## Data Availability

All data produced in the present work are contained in the manuscript and are available upon reasonable request to the authors

## Declarations

### Ethics approval and consent to participate

Written informed consent through patient consent form was taken from all study participants after reading the study information brochure. The study protocol had been approved by the institutional ethics committee of IPGME&R, Kolkata and have been performed in accordance with the ethical standards as laid down in ICMR Ethical Guidelines 2006.

### Consent for publication

Not applicable as no personal information is shared here.

### Availability of data and materials

All data generated or analysed during this study are included in this published article. The details are available from the corresponding author on reasonable request.

### Author contributions

A. Guin and A. Ghosh designed and conceptualised the study. A. Guin, SM, DB and SC all contributed in experiments and collection of data. A. Guin drafted the manuscript and all the authors edited and approved the manuscript.

## Acknowledgements

The authors are immensely thankful to Dr. Sumit Chakraborty, Assistant Professor, Department of Radiology, Institute of Postgraduate Medical Education and Research (IPGME&R), SSKM Hospital, Kolkata, India for radiological assistance. The authors also wish to thank the doctors, co-researchers, and the laboratory technicians of the department of Rheumatology, Institute of Postgraduate Medical Education and Research (IPGME&R), SSKM Hospital, Kolkata, India for patient recruitment and technical help.

