## Supplementary figures and images for "Complementary Use of Resveratrol Improves Low-Grade Chronic Inflammation Unresolved by Standard DMARD Therapy in Rheumatoid Arthritis"

### the original blots for western blot data of fig 3

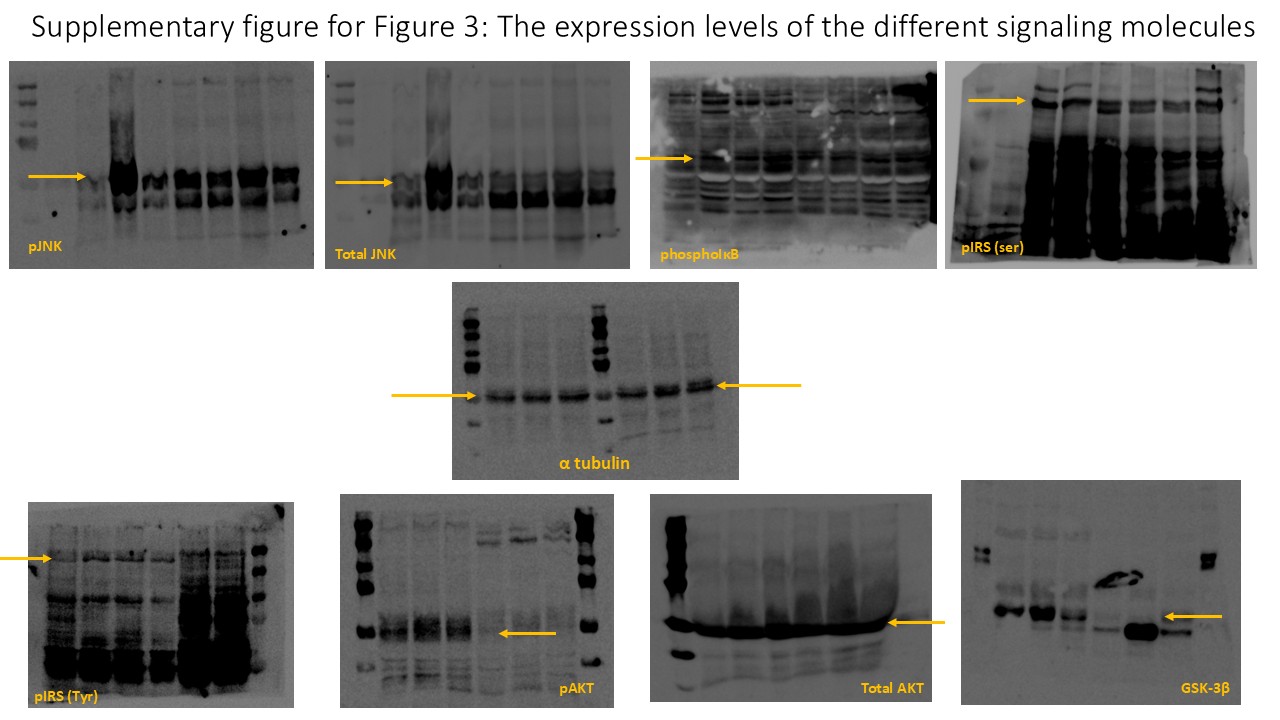

### the original blots for western blot for figure 5b

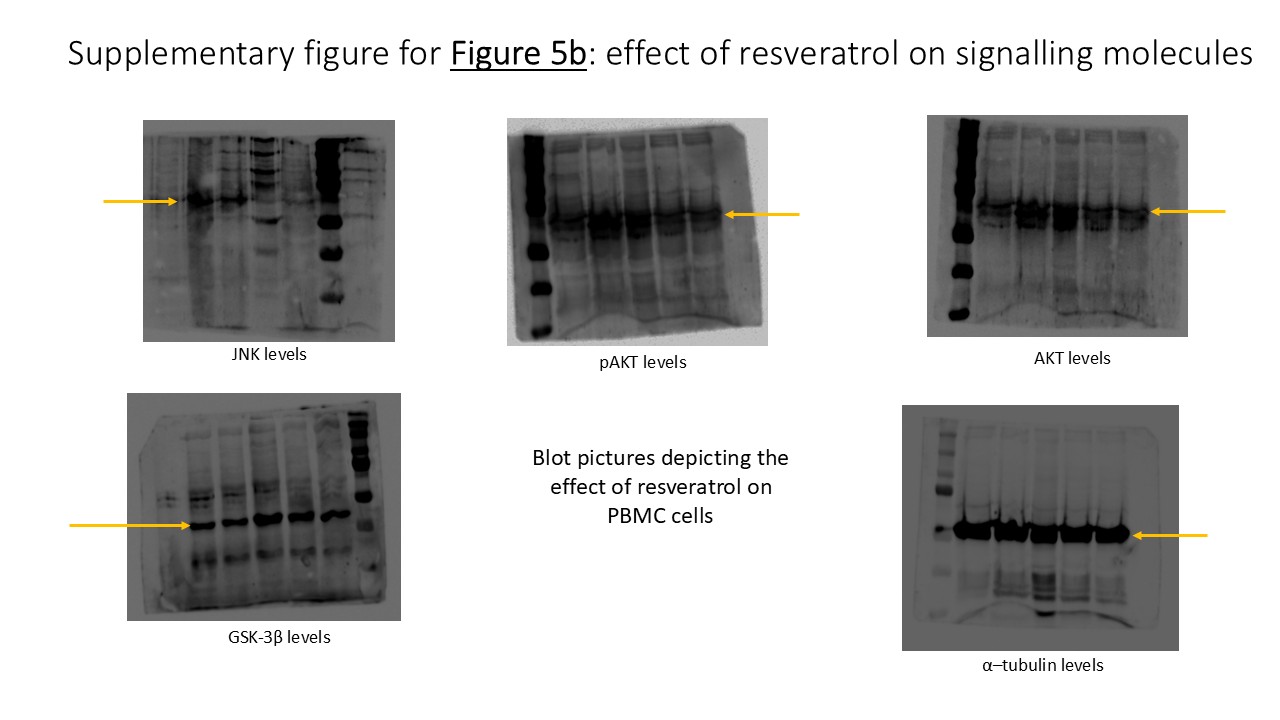
